# A normative torque-profile approach for quantifying elbow and wrist flexor hyper-resistance after stroke: a proof-of-principle study

**DOI:** 10.64898/2026.10.08.26364758

**Authors:** Sophie Julliand, Davy Laroche, Delphine Besson, Isabelle Fournel, Louis Riglet, Charalambos Papaxanthis, Jérémie Gaveau

## Abstract

**Background:** Post-stroke spasticity is commonly assessed using the Modified Ashworth Scale (MAS), despite its limited objectivity and variable reliability. This study introduces Score_E_, a novel isokinetic dynamometry-derived metric designed to quantify elbow and wrist flexor hyper-resistance by comparing patients’ torque-angle profiles with a healthy reference corridor.

**Methods:** Thirty-five post-stroke patients and twenty-one healthy controls were included. Elbow and wrist hyper-resistance was assessed using the MAS and an isokinetic dynamometer in two testing conditions: seated on the dynamometer chair (ergo chair) and seated in the patient’s wheelchair. Two dynamometer-derived metrics were analyzed: Score_E_ and peak torque (Peakτ). Feasibility, inter-rater and test-retest reliability, and associations with MAS grades were assessed. Exploratory PRE-POST changes following a 10-min local vibration session were also described.

**Results:** Score_E_ showed good to excellent inter-rater and test-retest reliability across joints and testing conditions. Inter-rater ICCs ranged from 0.81 to 0.89, and test-retest ICCs from 0.76 to 0.93. Peakτ showed moderate to good reliability. In contrast, MAS agreement was lower and more variable. Score_E_ showed moderate associations with MAS grades, particularly at the elbow, with weaker and less consistent associations at the wrist. MAS assessment was faster, but all assessments were well tolerated, with median pain ratings of 0/10 in all testing conditions. PRE-POST vibration changes were small and heterogeneous, preventing formal assessment of responsiveness.

**Conclusion:** Score_E_ provides a reliable, objective, and visually interpretable quantification of post-stroke elbow and wrist flexor hyper-resistance. By comparing the full torque-angle profile with normative data, this approach preserves information on both the magnitude and angular span of abnormal resistance, supporting its potential for targeted longitudinal monitoring.

## Introduction

Spasticity is a common and clinically relevant consequence of stroke, contributing to pain, functional limitations, difficulties in care, and increased burden of rehabilitation affecting individuals worldwide with various neurological disorders (1,2). Its prevalence after stroke is estimated at approximately 25%, with disabling spasticity affecting a smaller but clinically important proportion of patients (2). Accurate assessment is therefore essential for monitoring disease evolution, guiding therapeutic decisions, and evaluating interventions (3,4).

Spasticity has traditionally been defined as a velocity-dependent increase in stretch reflex activity (5). More recently, the Spasticity X Working Group proposed an updated consensus definition framing spasticity as a disorder of sensorimotor control following upper motor neuron disease, characterized by velocity and length-dependent involuntary muscle overactivity during passive stretch (6). Importantly, however, the resistance encountered during passive movement is multifactorial and may reflect both neural and non-neural mechanisms (7,8). Neural contributors include stretch hyperreflexia and involuntary background muscle activation, whereas non-neural contributors include altered muscle-tendon stiffness and connective tissue changes (5,8,9). For this reason, recent consensus recommendations encourage the broader term “hyper-resistance” to describe the overall abnormal resistance to passive stretch, and emphasize the need for objective parameters derived from instrumented assessment to distinguish its underlying components (8).

In clinical practice, the Modified Ashworth Scale (MAS) remains the most widely used tool for assessing spasticity because it is fast, simple and requires no equipment (10). However, the MAS provides only an ordinal rating of perceived resistance during manual passive movement. Its score depends on the examiner’s perception of resistance, the manually imposed stretch velocity, the tested range of motion, the patient’s position and the interpretation of categories (11–13).

Consequently, MAS reliability varies across joints, muscles, and raters, and the scale cannot quantify resistance in physical units or provide a continuous description of the resistive torque across a specific range of motion (10,11). These limitations are particularly problematic when subtle changes must be monitored longitudinally or when treatment effects must be evaluated in clinical trials.

To overcome these limitations, instrumented approaches have been developed to quantify the mechanical response to passive stretch. Isokinetic dynamometry is particularly relevant because it provides standardized angular velocities and simultaneous torque-angle recordings. Previous studies have used dynamometer-derived metrics such as peak resistive torque, mean torque, or threshold angle to characterize passive hyper-resistance and to compare instrumented measurements with clinical scales (14–18). These approaches improve standardization compared with manual assessment. Nevertheless, many existing metrics reduce the entire torque-angle profile to a single global value, such as peak torque. This may overlook where within the range of motion abnormal resistance occurs and may fail to distinguish a brief localized torque increase from a broader abnormal response over a larger portion of the movement.

A further challenge is that passive resistance is present even in healthy individuals and may also vary with joint, side, testing position, and movement velocity (8,19,20). Therefore, interpreting a patient’s torque profile requires a meaningful reference. Normative datasets and reference-based approaches have already shown value in instrumented assessment of passive hyper-resistance, as illustrated by NeuroFlexor normative data and cut-off values derived from healthy participants (20). However, few approaches directly compare the full torque-angle profile of post-stroke patients with a pointwise reference corridor derived from healthy participants while preserving information about both the magnitude and angular extent of the abnormal response. This remains an important gap, especially as recent reviews emphasize the need for more standardized, interpretable, and psychometrically validated technology-assisted measures of spasticity (4).

In this proof-of-principle study, we developed a normative torque-profile approach to quantify elbow and wrist flexor hyper-resistance after stroke using an isokinetic dynamometer. The proposed metric, Score_E_, quantifies torque excess outside a healthy reference corridor over all abnormal range of the passive stretch. The profile-based approach provides a continuous and visually interpretable representation of abnormal resistance, while also allowing the onset and amplitude of the abnormal response to be identified. We focused on elbow and wrist flexors because these muscle groups are commonly affected after stroke and are clinically relevant targets for spasticity management (21,22). The objectives of this study were to evaluate the feasibility and reliability of this dynamometer-based approach, to compare Score_E_ with a conventional torque-based metric, peak torque, and to assess its association with the MAS as the most widely used clinical comparator. In addition, exploratory PRE– POST changes following a short local muscle vibration session were described.

## Materials and Methods

### Population

Thirty-five patients from Dijon University Hospital were included in the study (Table 1), from July 2022 to October 2025. Two participants dropped out after visit 1, due to health-related reasons, leaving 33 analyzable patients at V2. All participants had experienced a stroke more than 3 months prior to evaluation. They all exhibited increased resistance to passive movement of the wrist and/or elbow flexors of the paretic arm, rated from 1 to 3 on the Modified Ashworth Scale (MAS). Additionally, twenty-one healthy participants were recruited to establish a reference baseline (Table 1).

**Table 1:** Participant characteristics. Values are presented as median [Q1-Q3], unless otherwise specified.

|  | Patients | Healthy participants |
| --- | --- | --- |
| Number | 35 | 21 |
| Gender | 11 females | 11 females |
| Age (years) | 67.0 [53.5-74.5] | 26.0 [24.0-28.0] |
| Time since stroke (years) | 1.9 [0.8-6.5] |  |
| Paretic arm | 17 Left / 18 Right |  |
| MAS Elbow Flexors (n = 34) | 1.25 [1.0-2.0] |  |
| MAS Wrist Flexors (n = 33) | 2.0 [1.0-2.0] |  |

All participants were screened to ensure the absence of other neurological, psychological, or musculoskeletal disorders, and reported pain levels ≤ 5 / 10 on the visual analog scale during elbow and wrist mobilization, to avoid interference with the study outcomes. The protocol was approved by the CPP Ile de France IV (number 2021-A03219-32, and ClinicalTrials NCT05315726). All participants provided informed consent, in accordance with the 2013 revision of the Declaration of Helsinki.

### General experimental setup

Patients attended two study visits, during which hyper-resistance of the wrist and elbow flexors was assessed using two different methods: an isokinetic dynamometer and a manual evaluation by a clinician (Figure 1). During the first visit (V1), inter-rater reliability was evaluated with all assessments conducted by two examiners. During the second visit (V2), test-retest reliability was assessed before vibration, and exploratory PRE-POST changes were described following a 10-minute vibration session applied to the arm. The order of the tests was randomized upon inclusion and remained consistent between V1 and V2, with an interval of 11 to 18 days separating the two visits. At V1, the order of the two examiners was also randomized, with one of them performing the assessments at V2. Both visits were conducted at the same time of day for each patient to minimize intra-participant circadian variations and fluctuations in passive resistance (23,24). Participants were instructed to maintain their usual lifestyle during the protocol period and advised not to start or modify any medication or treatment (e.g., botulinum toxin).

**Figure 1:**
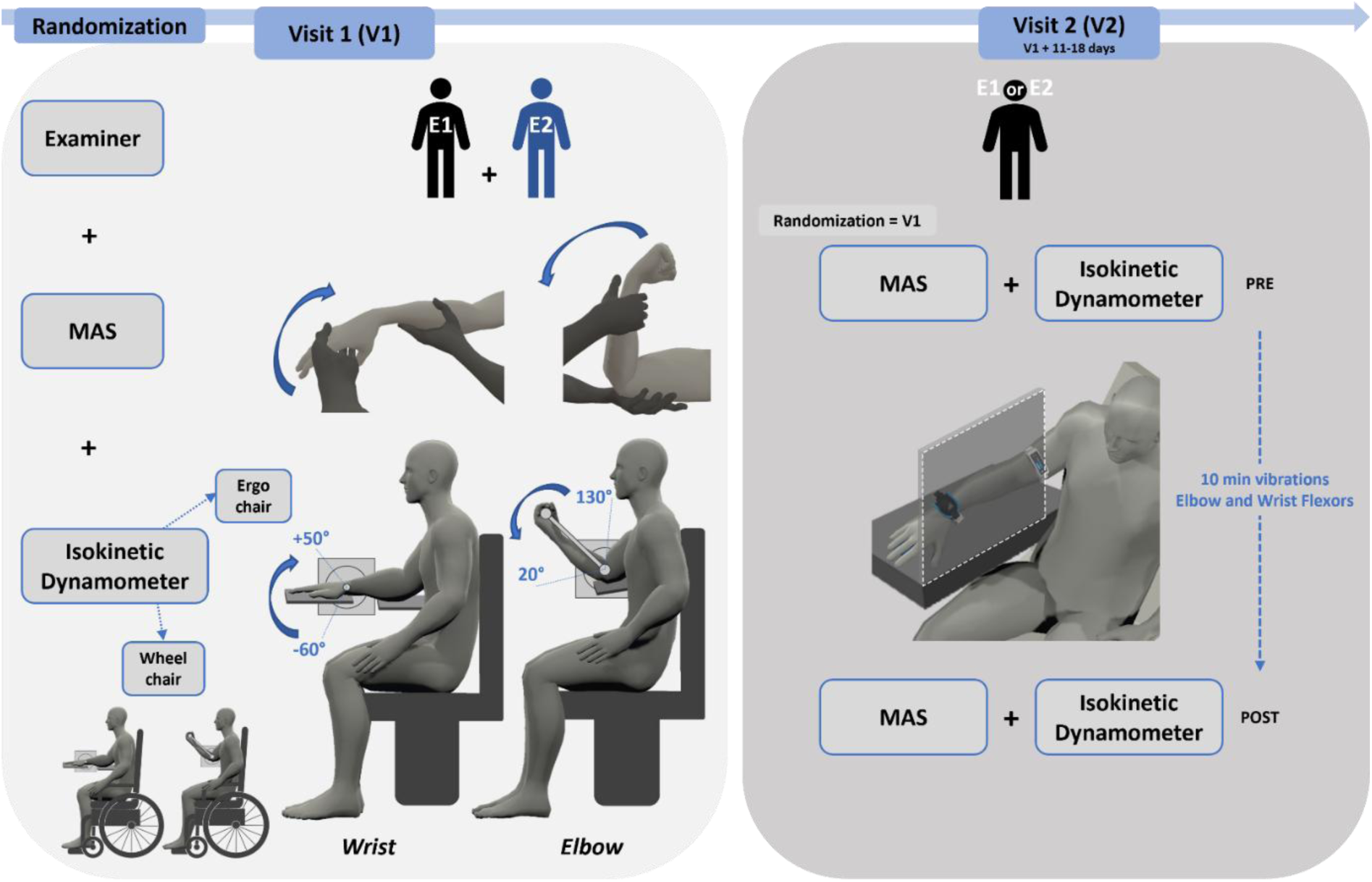
Schematic design of the study. Elbow and wrist flexors assessments were performed during two visits. During Visit 1 (V1), inter-rater reliability was assessed by two examiners (E1 and E2) using both the Modified Ashworth Scale (MAS) and the isokinetic dynamometer. Dynamometer assessments were performed in two testing conditions: seated on the ergo chair and seated in the participant’s wheelchair. During Visit 2 (V2), one of the examiners involved at V1 repeated the same assessments before (PRE) and after (POST) a 10-minute vibration session applied to the paretic arm. The order of examiners and assessments was randomized at inclusion.

Healthy participants underwent a single dynamometer assessment session using the same dynamometer protocol. Both upper limbs were assessed at the elbow and wrist in both testing conditions (ergo chair and wheelchair), to provide side-, joint-, and condition-specific normative reference data.

### Hyper-resistance assessments

#### Isokinetic dynamometer

Passive resistance of the elbow and wrist flexors was measured using an isokinetic dynamometer (Biodex System 4, Biodex Medical Systems inc., Shirley, USA). The dynamometer’s signals were converted from analog to digital at a frequency of 2000 Hz using the Biopac acquisition system (AcqKnowledge, Biopac System Inc. Goleta, CA, USA). During testing, patients were seated comfortably on the ergo chair or in their wheelchair, with their trunk securely strapped to minimize compensatory movements. For all assessments, the dynamometer module was tilted at 0° and rotated 10° outward (Figure 2).

**Figure 2:** Participant positioning during isokinetic dynamometer assessments, of the Wrist (on the left) and Elbow (on the right).

Adjustments were made to ensure maximum relaxation of the assessed limb and to achieve the following standardized positions. For the evaluations on the designated ergo chair, i) for the elbow flexors assessment, the elbow was supported on an armrest with the hand supinated and strapped around the handle (Figure 2). The shoulder was positioned at approximately 30° of abduction, flexion, and external rotation, with the trunk-to-hip angle at around 100°; ii) for wrist flexors assessment, the elbow and forearm were placed on a specific armrest designed to keep the forearm horizontal. The shoulder and trunk positioning mirrored that used during elbow evaluation, with the elbow maintained at about 80°. The hand was firmly strapped to a support specifically designed for the experiment (Figure 2). The rotation axis of the machine was aligned with the rotation axis of the elbow or wrist. For the evaluations performed in the wheelchair, positions were identical but without armrest support.

The range of motion was standardized to 110° for all patients and assessments, with specific angular limits set from 130° to 20° of flexion for the elbow (0° being the elbow fully extended) and from -60° of flexion to 50° of extension for the wrist (0° being the neutral position of the wrist). Following two practice trials at 30°/s, which allowed participants to express any discomfort and familiarize themselves with the movement of the machine, the formal evaluation was conducted at an angular velocity of 180°/s (17). Two trials were recorded, with a 30-second rest period between them. During the evaluation, the machine passively moved the joint through its range of motion, while patients were instructed to fully relax.

During V1, the testing procedure was performed independently by two different examiners, who were responsible for positioning the patient on the machine, performing the two practice trials, and carrying out the evaluation. During V2, the procedure was completed by only one examiner.

#### Modified Ashworth Scale

Clinical resistance to passive movement of the elbow and wrist flexors was evaluated using the Modified Ashworth Scale (MAS), a standardized clinical scale commonly used as a surrogate measure of spasticity. Clinicians subjectively rate the resistance across 6 levels, ranging from 0 (no increase in muscle tone) to 4 (rigid joint), as detailed in Table 2 (25). Each examiner performed passive mobilization of the joint to assess the resistance encountered during stretching of the targeted muscle group. Participants were seated comfortably in a chair or wheelchair, in a position similar to that used on the dynamometer, as resistance to passive movement can vary depending on body positioning (26). The assessment followed these steps: The examiner first moved the joint slowly to determine the available range of motion and detect any discomfort. Then, two rapid passive stretches were applied to the joint, with a 30-second interval between each stretch. The clinician assigned a MAS score for each trial.

**Table 2:** Modified Ashworth Scale definitions. MAS scores were defined according to Bohannon and Smith (1987) (25). ROM: range of motion.

| Score | Description |
| --- | --- |
| 0 | No increase in tone |
| 1 | Slight increase in tone. Catch and release or minimal resistance at the end of the ROM. |
| 1 + | Slight increase in tone. Catch and release or minimal resistance throughout the remainder (less than half) of the ROM. |
| 2 | More marked increase through most of the ROM, but affected part moved easily. |
| 3 | Considerable increase in tone, passive movement difficult. |
| 4 | Affected part rigid in flexion or extension. |

During V1, the testing procedure was performed independently by two different examiners, whereas during V2, it was conducted by only one examiner.

### Feasibility evaluations

To assess the practicality of the two evaluation methods (MAS and isokinetic dynamometer), the duration of each procedure performed by each examiner was timed during V1. For dynamometer assessments (ergo chair or wheelchair), timing included patient positioning, setup, and the complete elbow and wrist assessment. In the ergo-chair condition, transfer to and from the dynamometer chair was included. For the MAS assessment, the timer started when the clinician was instructed to start the procedure, and stopped at the end of the assessment.

Patient pain levels were evaluated using a visual analog scale (VAS), ranging from 0 (no pain) to 10 (worst pain ever experienced). Pain ratings were recorded for each evaluation and examiner during V1 to monitor procedure-related discomfort.

### Vibrations (V2 only)

During V2, a 10-minute continuous mechanical vibration session applied to the paretic limb was used to explore short term PRE-POST changes in MAS and dynamometer-derived outcomes. This duration was chosen based on previous studies demonstrating its effectiveness in modulating spasticity (27). The vibration module (Vibramoov, Technoconcept, Manosque, France) was placed directly on the skin, positioned approximately 2 cm proximal to the elbow crease, and 2 cm proximal to the wrist crease. To ensure secure placement and consistent pressure, the vibrators were fastened around the arm and forearm using an adjustable strap (Figure 1). Individual adjustments were made to optimize comfort and fit for each patient.

Vibrations were continuously administered for 10 minutes at a frequency of 80 Hz and an amplitude of 2 mm (according to the manufacturer’s specifications). To minimize the influence of visual feedback and reduce activation of the tonic vibratory reflex, the patient’s forearm was obstructed from view using a cardboard box (28,29). During vibrations, patients were comfortably seated in a semi-reclined posture to promote muscle relaxation. Their arm was supported at the elbow and wrist using malleable cushions, with the elbow semi-flexed, pronated, and the wrist in a neutral position. Patients were instructed to remain relaxed and focus on the vibratory sensations.

### Data analysis

#### Isokinetic dynamometer

A custom-made script in MATLAB (MATLAB R2024a; MathWorks, Natick, MA) was developed to process all data recorded by the isokinetic dynamometer. First, torque and position signals were low-pass filtered using a third-order Butterworth filter with a 10Hz cut-off frequency. Angular velocity was then obtained by numerical differentiation of the position signal. Movement onset and offset were defined as the first and last samples at which angular velocity exceeded 5% of the peak angular velocity of that trial. The target velocity of 180°s^-1^ was reached within approximately 200ms, and the average movement duration was 1.09 seconds. For each trial, torque and position signals were time-normalized and linearly interpolated to 2000 points. The same fixed 2000-point representation was used for all participants, sessions, joints, and testing conditions to ensure identical discretization for Score_E_ computation. The two recorded trials of the patient were processed separately and then averaged for each examiner, joint and testing condition before statistical analysis.

Torque signals were analyzed as recorded from the acquisition system, without gravity correction or normalization to limb segment mass, as limb anthropometrics were not measured. To reduce differences related to initial torque offsets between patients and testing sessions, each patient torque profile was shifted so that its initial value matched the center of the corresponding reference corridor at movement onset. Torque sign was not normalized across sides; therefore, right and left limbs were analyzed using their original torque direction. Each patient profile was compared with the corresponding reference corridor built from the same joint, testing condition, and side/direction.

This reference corridor was built separately for each joint, testing condition, and side using the corresponding limb data from the 21 healthy control participants (the grey area in Figure 3). As all healthy participants were assessed bilaterally in both testing conditions, each side-specific reference corridor was derived from the corresponding side of the healthy sample. Control data were processed following the same procedure as patient data described above. At each normalized sample, the mean and standard deviation (SD) were computed, and the reference corridor was defined as the pointwise mean ± 1 SD. Patient torque values exceeding either the upper or lower boundary of the reference corridor (depending on the joint/side/condition) were considered outside the reference range. If several abnormal segments were present within the same trial, all abnormal samples were included in the calculation of Score_E_. For each abnormal sample, torque excess was computed relative to the nearest boundary of the reference corridor (Figure 3). Score_E_ was then calculated as the discrete Euclidean norm of these torque excess values across all abnormal samples (Eq. 1).

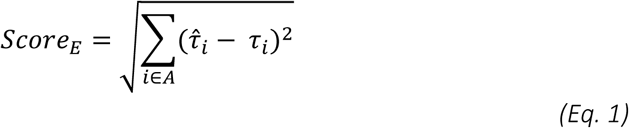

**Figure 3:**
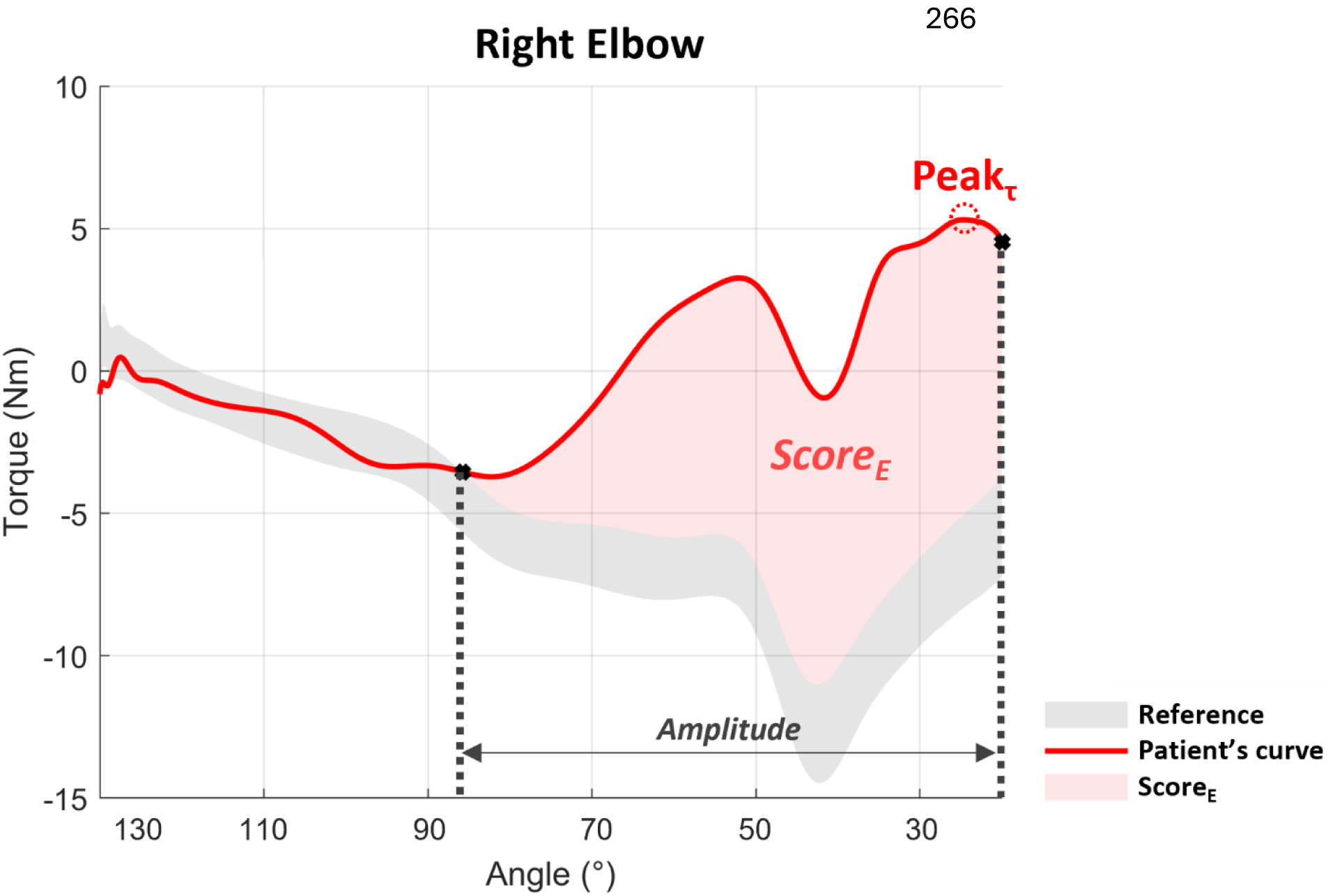
Representative computation of Score_E_ from the torque profile. Example of a torque-angle profile obtained during a passive elbow extension in one post-stroke participant. The gray area depicts the reference corridor derived from healthy participants. The red curve represents the patient’s torque profile. The shaded red area indicates samples exceeding the reference corridor and contributing to Score_E_. The onset and end angles correspond to the first and last angular positions at which the patient profile exceeded the reference corridor. Peak_Ƭ_ corresponds to the highest Torque absolute value throughout the entire ROM. In this example, Score_E_ was 298.9 N.m, with an onset angle of catch at 85.8°.

*A represents the set of all abnormal samples (i.e., all normalized samples for which the patient torque curve exceeded the reference corridor). For each abnormal sample i, τ_i_ is the patient torque value and 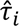 is the nearest boundary of the reference corridor. If several abnormal segments were present, all abnormal samples were included in the calculation*.

To compare with the previous results from the existing literature, we also calculated Peak Torque (Peak_Ƭ_) in N.m (Eq. 2), corresponding to the maximal absolute value of the patient’s torque curve (see Figure 3).

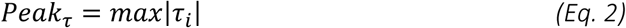

*with τ_i_ representing the patient’s Torque values*

#### Modified Ashworth Scale

The highest score of the two trials for each examiner during V1 and during the PRE and POST evaluations during V2 was retained for each muscle group. To facilitate interpretation, the MAS score of 1+ was assigned a value of 1.5 to preserve its ordinal position between grades 1 and 2 (30).

### Statistical analysis

Statistical analyses were conducted using JASP (version 0.95.3; University of Amsterdam, The Netherlands). Data distribution was assessed using the Shapiro-Wilk test. Statistical significance was set at *p ≤ 0.05*. When the mean of the two V1 examiner ratings was converted back into a MAS category, predefined intervals were used: 0 for mean score of 0; 1 for values from 0.5 to 1; 1+ for values from 1.25 to 1.75; 2 for values from 2 to 2.5; and 3 for a mean score of 3.

#### Feasibility of the three methods (i.e., MAS, isokinetic on ergo chair and on wheelchair)

The mean duration across the two examiners during V1 was calculated for the three assessments. For pain, a single VAS score was calculated for each participant and each assessment condition during V1: when both wrist and elbow VAS scores were available, their mean was used; when only one joint was assessed, the available VAS score was retained. A Shapiro-Wilk test revealed that neither duration nor pain data were normally distributed across the three conditions. A Friedman test was performed, with Conover’s post-hoc comparisons if the results were significant. Effect sizes were calculated using Kendall’s W, interpreted as small (<0.3), moderate (0.3-0.5), or large (>0.5).

#### Reliability and agreement of the two methods

The intraclass correlation coefficient (ICC) was employed to assess the reliability of isokinetic dynamometer-derived metrics (Score_E_ and Peak_Ƭ_). Inter-rater reliability, corresponding to the agreement between the two examiners during V1, was evaluated using a two-way random effects model for single measurements and absolute agreement, reported as ICC (2,1). This model was chosen because the two examiners were considered representative of trained raters to whom the results may generalize.

Test-retest reliability was assessed between V1 and the PRE assessment of V2. Because the examiner who performed the V2 assessments was one of the two examiners involved at V1, the mean of the two examiner-specific values obtained during V1 was used as the V1 reference value for ScoreE and PeakT, and compared with the value obtained at V2 PRE. Test-retest reliability was evaluated using a two-way mixed-effects model for single measurements and absolute agreement, reported as ICC(3,1) (31). For both analyses, each dynamometer-derived value corresponded to the average of the two recorded trials. Bland-Altman plots were used to identify systematic bias and individual differences between measurements (Altman & Bland, 1983 ; Supplementals Figure 5).

For the MAS, inter-rater agreement was assessed between the two examiners at V1 using linearly weighted Cohen’s kappa (k) (33). For test-retest agreement, the mean of the two V1 examiner ratings was first converted back into a MAS category using the predefined grouping rule described above and was then compared with the MAS grade obtained at V2 PRE, also using linearly weighted Cohen’s kappa. The ICC values were interpreted as follows (31): < 0.50 indicated poor reliability, 0.50 – 0.75 signified moderate reliability, 0.76 – 0.90 represented good reliability, and > 0.90 indicated excellent reliability. Similarly, kappa values were categorized as follows: ≤ 0.20 for slight, 0.21 – 0.40 for fair, 0.41 – 0.60 for moderate, 0.61 – 0.80 for substantial and > 0.80 for almost perfect agreement (34).

#### Association between MAS and dynamometer-derived metrics

Associations between MAS scores and dynamometer-derived metrics were assessed using two-sided Kendall’s tau-b correlation coefficient (τb), because MAS is an ordinal scale with tied ranks (35). For each participant, examiner-specific V1 values were averaged across the two examiners before correlation analyses, for both dynamometer-derived metrics and MAS. For MAS, the mean of the two numerically coded examiner-specific scores was retained as the participant-level V1 value for the correlation analyses. Correlations were calculated between MAS and Score_E_, as well as between MAS and Peak_Ƭ_. P-values were adjusted for multiple comparisons across the four MAS-dynamometer associations per joint, using the Holm–Bonferroni correction method (36).

For descriptive analyses by MAS category, the same predefined grouping rule described above was applied to the mean V1 MAS score to retain a clinically interpretable presentation. Associations between Score_E_ values obtained in the ergo-chair and wheelchair conditions were additionally assessed using Spearman’s rank correlation coefficient (rho), with 95% confidence intervals.

#### Exploratory PRE-POST changes of the two methods after vibrations

Changes following the 10-minute vibration session were analyzed descriptively. For Score_E_, individual changes were calculated as POST minus PRE values (ΔPrePost) for each joint and testing condition and summarized using median changes and interquartile ranges. For the MAS, changes were described categorically as the number of participants showing a decrease, no change, or increase in MAS grade. Because the vibration protocol was not expected to provide a standardized therapeutic intervention and did not induce a sufficiently large or consistent change in hyper-resistance, this analysis was considered exploratory and was not used to formally assess responsiveness.

No imputation was performed, analyses were conducted on available cases.

## Results

### Feasibility

Assessment duration (elbow plus wrist) was recorded during V1 for both examiners, along with patient’s perceived pain during each assessment. The assessment duration differed significantly across the three conditions (p < 0.01, W = 0.907). The dynamometer assessment performed on the ergo chair (including patient transfers) was the longest procedure, with a median duration of 15 minutes 59s, whereas MAS assessment was substantially shorter, with a median duration of 2 min 03s. Post-hoc comparisons indicated that all three conditions differed significantly from each other (Figure 4A). Pain ratings remained low across all procedures. Median VAS were 0.0/10.0 in all conditions, and no significant difference was observed between methods (Figure 4B).

**Figure 4:**
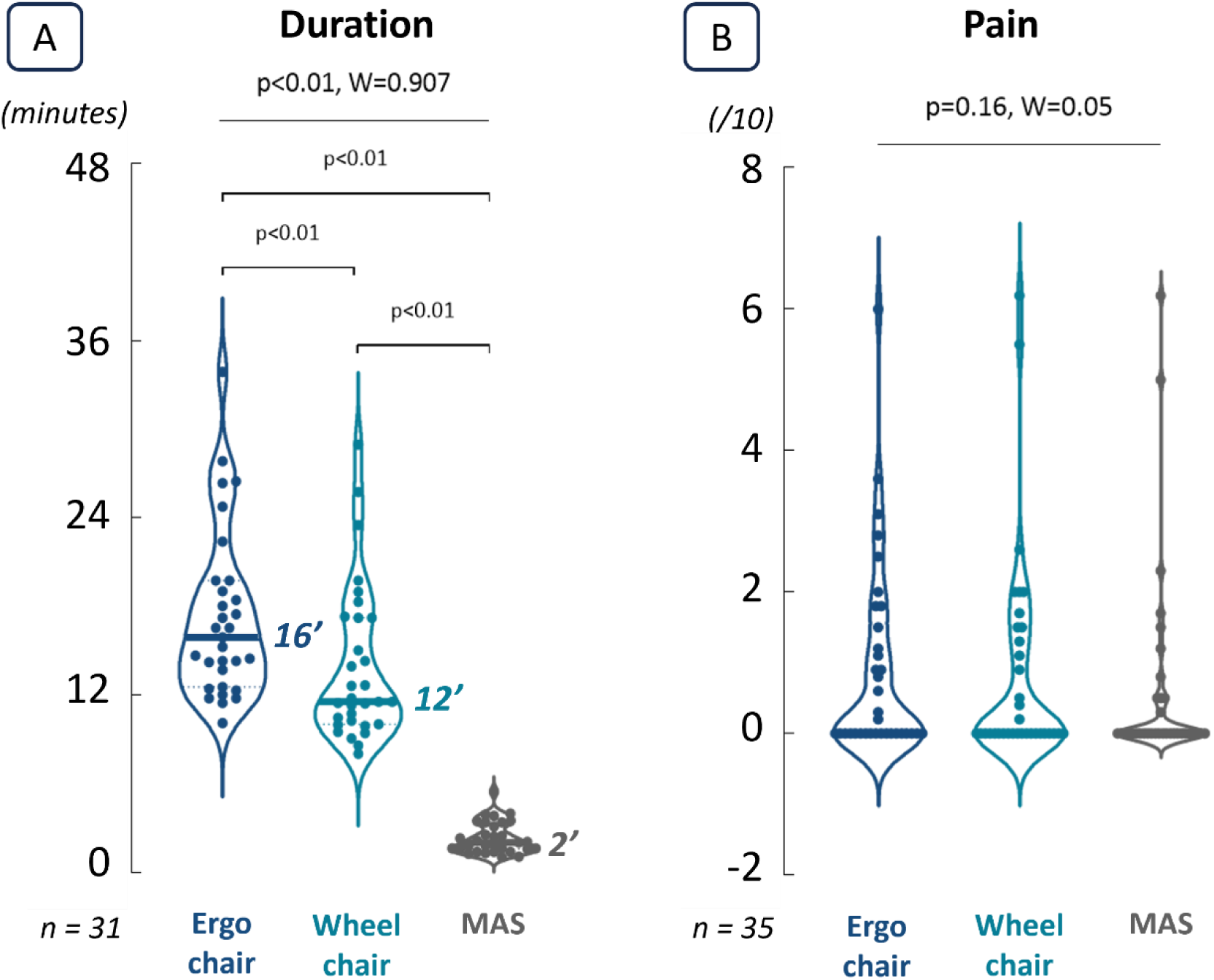
Feasibility of the assessment procedures. (A) Distribution of assessment duration across the three conditions at V1: ergo chair, wheelchair, and MAS. Duration (in minutes) included both elbow and wrist assessments. (B) Pain intensity reported by patients during each assessment condition, measured using a 10-point visual analog scale (VAS). In both panels, dots represent individual patients and horizontal lines indicate medians. Global differences across the three conditions were assessed using Friedman tests, with Kendall’s W reported as effect size. When significant, pairwise Conover tests were performed, and p-values are displayed above the corresponding comparisons.

### Reliability and agreement

Table 3 presents the inter-rater and test-retest reliability of the clinical and dynamometer-based assessments for the elbow and wrist flexor. For the dynamometer-derived outcomes, inter-rater reliability corresponded to the agreement between the two examiners during V1, whereas test-retest reliability corresponded to the comparison between the mean of the two examiner-specific values at V1 and the PRE assessment of V2.

**Table 3:**
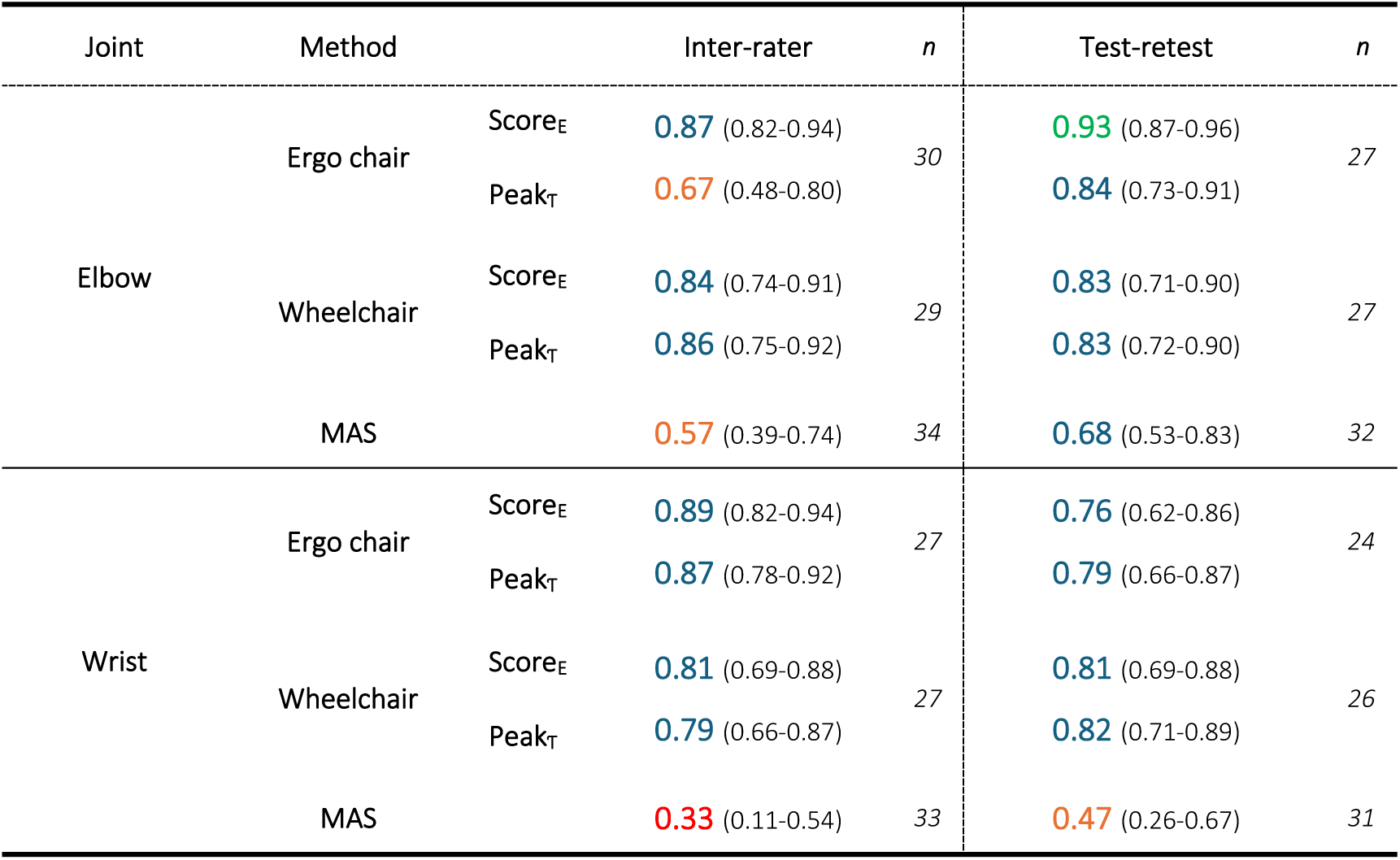
Inter-rater and test-retest reliability of MAS and dynamometer-derived metrics. For dynamometer-derived metrics, values correspond to single-measure intraclass correlation coefficient (ICC) with 95% confidence intervals. For MAS, values correspond to linearly weighted Cohen’s kappa coefficients (k) with 95% confidence intervals. The number of patients included in each analysis is indicated as n. Green indicates excellent agreement, blue for good/substantial, orange for moderate, and red for poor/fair.

| Joint | Method |  | Inter-rater | *n* | Test-retest | *n* |
| --- | --- | --- | --- | --- | --- | --- |
| Elbow | Ergo chair | Score<sub>E</sub> | 0.87 (0.82-0.94) | 30 | 0.93 (0.87-0.96) | 27 |
|  |  | Peak<sub>T</sub> | 0.67 (0.48-0.80) |  | 0.84 (0.73-0.91) |  |
|  | Wheelchair | Score<sub>E</sub> | 0.84 (0.74-0.91) | 29 | 0.83 (0.71-0.90) | 27 |
|  |  | Peak<sub>T</sub> | 0.86 (0.75-0.92) |  | 0.83 (0.72-0.90) |  |
|  | MAS |  | 0.57 (0.39-0.74) | 34 | 0.68 (0.53-0.83) | 32 |
| Wrist | Ergo chair | Score<sub>E</sub> | 0.89 (0.82-0.94) | 27 | 0.76 (0.62-0.86) | 24 |
|  |  | Peak<sub>T</sub> | 0.87 (0.78-0.92) |  | 0.79 (0.66-0.87) |  |
|  | Wheelchair | Score<sub>E</sub> | 0.81 (0.69-0.88) | 27 | 0.81 (0.69-0.88) | 26 |
|  |  | Peak<sub>T</sub> | 0.79 (0.66-0.87) |  | 0.82 (0.71-0.89) |  |
|  | MAS |  | 0.33 (0.11-0.54) | 33 | 0.47 (0.26-0.67) | 31 |

Overall, dynamometer-derived metrics showed moderate to excellent reliability across joints and testing conditions. For Score_E_, inter-rater reliability was good for both the elbow and wrist, with ICC values ranging from 0.81 to 0.89 across testing conditions. Test-retest reliability was excellent for the elbow in the ergo chair condition and good in the remaining conditions, with ICC values ranging from 0.76 to 0.93. Peak_Ƭ_ showed good reliability in most conditions, although inter-rater reliability was only moderate for the elbow in the ergo chair condition.

By contrast, MAS agreement was consistently lower than the dynamometer-derived metrics. At the elbow, MAS showed moderate inter-rater and substantial test-retest agreement. At the wrist, inter-rater agreement was fair, whereas test-retest agreement was moderate.

Bland–Altman plots were used to visually inspect systematic bias and heteroscedasticity and are provided in Supplementary Figure 5. These analyses showed generally small mean biases for Score_E_ across joints and testing conditions, suggesting limited systematic disagreement between measurements. Inter-rater biases were close to zero for both the elbow and wrist. The limits of agreement were wider at the elbow than at the wrist. Test–retest comparisons showed a similar pattern, with relatively small mean biases but wider limits of agreement at the elbow. Visual inspection did not reveal a marked systematic bias.

### Association MAS-dynamometer

Table 4 presents the associations between MAS grades and dynamometer-derived metrics in all conditions, and Fig. 5 illustrates the distribution of Score_E_ across MAS grades. Representative torque-angle profiles illustrating Score_E_ across different MAS grades are provided in Supplementary Fig. 1.

**Table 4:** Associations between MAS and dynamometer-derived metrics. Kendall’s tau-b correlations (Ƭb) with 95% confidence intervals are reported for associations between MAS and ScoreE or Peak_Ƭ_ at the elbow and wrist, in the ergo chair and wheelchair conditions. P-values were adjusted for multiple comparisons using the Holm– Bonferroni procedure. Significant (p ≤ 0.05) adjusted p-values are shown in bold. Sample sizes were n = 31 for the elbow ergo chair condition, n = 29 for the elbow wheelchair condition, and n = 27 for both wrist conditions.

|  |  | Elbow |  | Wrist |  |
| --- | --- | --- | --- | --- | --- |
|  |  | p | Tb (95%CI) | p | Tb (95%CI) |
| $Score_E - MAS$ | Ergo chair | **0.004** | 0.489 (0.311; 0.666) | **0.020** | 0.409 (0.163; 0.655) |
|  | Wheelchair | **0.003** | 0.479 (0.248; 0.710) | 0.172 | 0.248 (-0.012; 0.507) |
| $Peak_T - MAS$ | Ergo chair | 0.072 | -0.235 (-0.533; 0.063) | **0.030** | 0.370 (0.126; 0.613) |
|  | Wheelchair | **0.002** | -0.474 (-0.697; -0.251) | 0.162 | 0.201 (-0.083; 0.486) |

**Figure 5:**
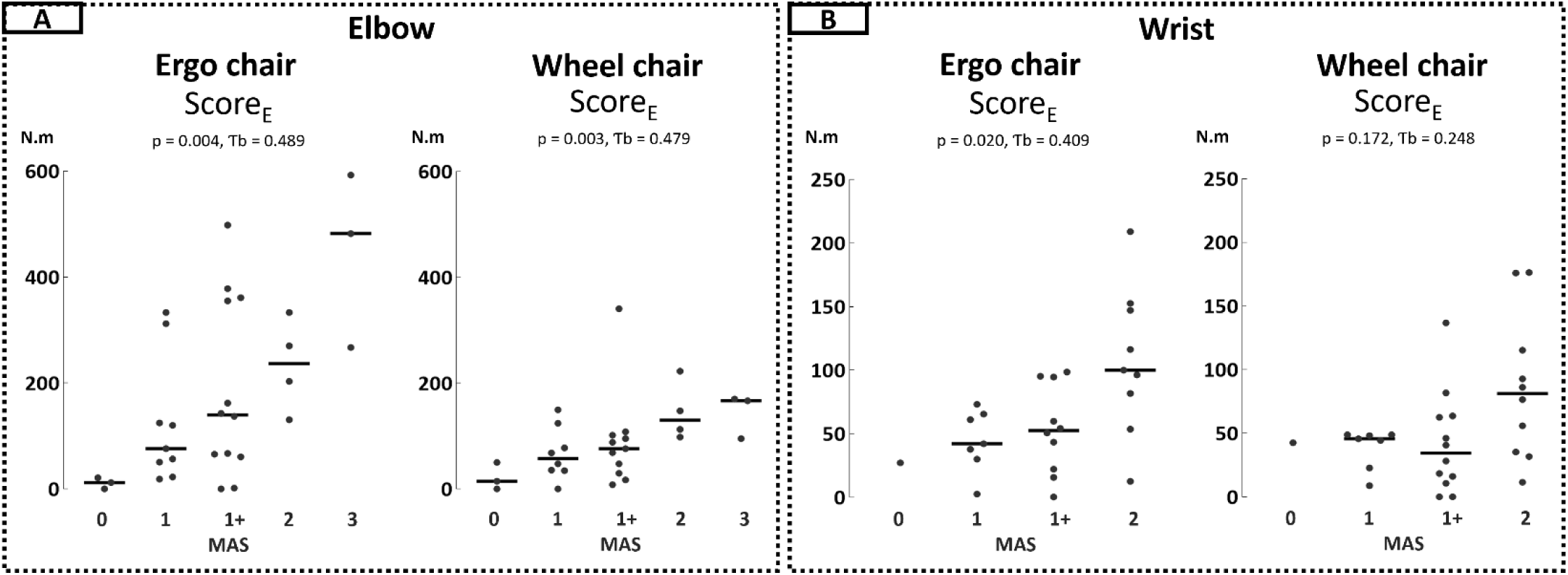
Distribution of Score_E_ according to MAS grade. Individual Score_E_ values are displayed according to MAS grade for elbow (A) and wrist (B) flexors, in the ergo chair and wheelchair conditions. Each dot represents one patient, and horizontal lines indicate median values within each MAS grade. Kendall’s tau-b coefficients and Holm-Bonferroni-adjusted p-values are reported for each condition.

At the elbow, Score_E_ showed moderate positive correlations with MAS in both testing conditions. Peak_Ƭ_ showed a significant negative association with MAS only in the wheelchair condition. At the wrist, correlations were weaker overall, with significant associations observed only in the ergo chair condition.

The corresponding distributions for Peak_Ƭ_ are provided in Supplementary Fig 3.

### Exploratory PRE-POST changes after vibration

Individual PRE–POST changes following the 10-minute vibration session are presented in Table 5 and Figure 6. Overall, Score_E_ changes were small, heterogeneous and bidirectional across participants, whereas MAS grades remained unchanged in most participants. Taken together, these findings indicate that the vibration session did not induce a consistent reduction in hyper-resistance at the group level. Because no minimal detectable change threshold was applied, these variations should be interpreted descriptively and not as evidence of responsiveness.

**Table 5:** Exploratory PRE-POST changes following the 10-minute vibration session. ΔPrePost values are reported as median [IQR]. Negative values indicate a numerical decrease after vibration, whereas positive values indicate numerical increase. For Score_E_, increase, decrease, and no change refer to the direction of the numerical POST–PRE difference. For MAS, increase, decrease, and no change refer to changes in MAS grade. No minimal detectable change threshold was applied; therefore, these results should be interpreted descriptively.

| Joint | Outcome | Condition | ΔPrePost | Increase | Decrease | No change |
| --- | --- | --- | --- | --- | --- | --- |
| Elbow | Score<sub>E</sub> | Ergo chair | -11.9 [67.4] | 9 (32%) | 18 (64%) | 1 (4%) |
|  |  | Wheelchair | -2.2 [55.8] | 12 (43%) | 15 (53%) | 1 (4%) |
|  | MAS |  | 0.0 [0.0] | 0 (0%) | 5 (16%) | 27 (84%) |
| Wrist | Score<sub>E</sub> | Ergo chair | -1.7 [32.2] | 10 (38%) | 14 (54%) | 2 (8%) |
|  |  | Wheelchair | 0.77 [14.1] | 13 (50%) | 11 (42%) | 2 (8%) |
|  | MAS |  | 0.0 [0.0] | 0 (0%) | 7 (23%) | 24 (77%) |

**Figure 6:**
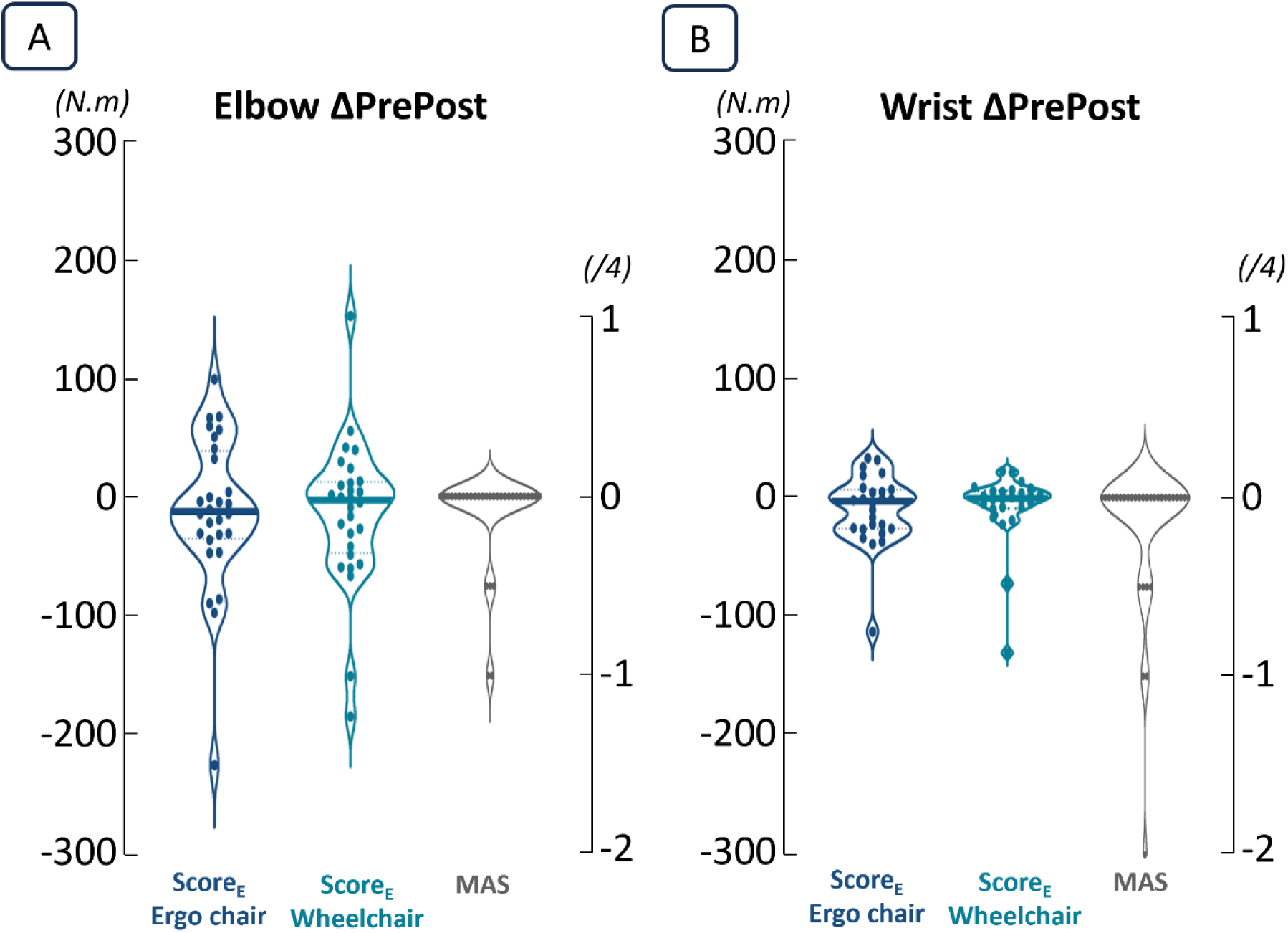
Exploratory individual PRE-POST changes following vibration. Violin plots show the distribution of individual ΔPrePost for Score_E_ and MAS at the elbow (A) and wrist (B). Dots represent individual patients and horizontal lines the median changes. Negative values indicate a decrease and positive values an increase. For MAS, grade 1+ was coded as 1.5 for calculation of ΔPrePost.

## Discussion

This proof-of-principle study investigated a new normative torque-profile approach for quantifying post-stroke upper-limb hyper-resistance using isokinetic dynamometry. The proposed metric, Score_E_, quantifies torque excess outside a healthy reference corridor derived from the full torque-angle profile during passive stretch. This approach was designed to provide a continuous and visually interpretable measure of abnormal resistance, while preserving information on the angular portion of the movement in which the abnormal response occurs.

The main findings were that Score_E_ showed good to excellent inter-rater and test-retest reliability for both elbow and wrist flexors, in both the ergo chair and wheelchair testing conditions, whereas MAS agreement was lower and more variable across joints. Score_E_ also showed moderate associations with MAS grades, with stronger and more consistent results at the elbow than at the wrist. Both assessment methods were well tolerated, although the dynamometer-based procedure required a longer administration time than the MAS.

Finally, exploratory PRE-POST changes were small and heterogeneous, indicating that this protocol did not induce a sufficiently consistent modification of hyper-resistance to formally assess responsiveness.

### Feasibility and potential clinical use

The widespread use of the MAS in clinical settings is largely due to its rapid administration, simplicity and ease of integration into routine neurological assessments (10). Our findings support this clinical advantage, as the MAS assessment took less than 4 minutes for both joints, whereas the dynamometer-based assessment required approximately 16 minutes in the ergo chair and 12 minutes in the wheelchair condition. However, this longer assessment time should be balanced against the greater standardization and richer quantitative information provided by the dynamometer-derived metrics, together with the good reliability observed particularly for Score_E_ in the present study. The MAS therefore remains more suitable for rapid screening, particularly when multiple joints need to be assessed during a routine clinical examination.

By contrast, isokinetic dynamometry may be more appropriate when a precise and reproducible follow-up of one or two clinically relevant joints is required. In clinical practice, such an approach could be useful for monitoring joints targeted by specific interventions, such as botulinum toxin injections, orthotic management, or focused rehabilitation programs. In research settings, the continuous torque-angle data provided by the dynamometer may be particularly valuable for characterizing treatment effects in greater detail than ordinal clinical scales, especially when changes are subtle or do not result in change in MAS grade (15,16). This positioning is consistent with current recommendations toward combining clinical examination with more standardized quantitative measures when a detailed characterization of hyper-resistance is required, rather than considering instrumented assessment as a replacement for bedside clinical evaluation (3,4).

The feasibility of the dynamometer-based assessment was also supported by its good tolerance, with median pain ratings of 0/10 in all testing conditions. Nevertheless, this result should be interpreted cautiously, as participants with severe pain during passive upper-limb mobilization were not included in the study. The procedure may therefore be less feasible in patients with painful shoulder conditions, severe contractures, or marked discomfort during passive positioning.

Finally, although isokinetic dynamometers such as the Biodex used in this study provide standardized and reliable measurements (37), they remain bulky, non-portable, and require dedicated space and trained personnel. This limits their use as routine bedside tools. Recent developments in portable and robotic devices, such as the NeuroFlexor for wrist and finger assessment (20) and robotic systems for elbow evaluation (16,38), reflect the growing need for more accessible instrumented assessments. In this context, the present normative torque-profile approach could potentially be transferred to more portable systems, provided that standardized passive mobilization and torque-angle recording can be achieved. The computational principle of Score_E_ is not specific to the Biodex and could therefore, in principle, be implemented on other devices capable of providing synchronized and sufficiently accurate torque and angular-position signals.

### Reliability of the dynamometer-derived metrics and MAS

The proposed Score_E_ approach exhibited good to excellent inter-rater and test-retest reliability for both elbow and wrist flexors, across the ergo chair and wheelchair testing conditions. This high reliability is consistent with previous studies showing that standardized passive mobilization using isokinetic or robotic devices can reduce examiner-dependent variability by controlling joint velocity, range of motion, and torque recording conditions (16–18,39,40). In particular, Sin et al. reported improved test-retest and inter-rater reliability for stretch-reflex-related measurements, including the angle of catch, when using an isokinetic device in post-stroke patients with mild to moderate elbow spasticity (16). The present findings extend this literature by showing that a reference-based analysis of the full torque-angle profile can also provide reproducible measurements of abnormal hyper-resistance.

Importantly, relative reliability should not be considered equivalent to absolute agreement. Previous work has shown that high ICC values may coexist with substantial individual measurement error. For example, Çetin et al. reported ICCs of 0.82–0.87 for wrist torque measurements but relatively large SEM and smallest real difference values (18). This distinction supports the complementary use of Bland– Altman analyses in the present study. Although mean biases were small, the wider limits of agreement observed at the elbow indicate that individual changes should be interpreted cautiously even when ICC reliability is good. Future studies should carefully determine the standard error of measurement and minimal detectable change of Score_E_ before it is used to classify individual longitudinal changes.

Peak_Ƭ_ also showed moderate to good reliability in most conditions, supporting the overall robustness of the dynamometer-based assessment. This is consistent with previous post-stroke studies reporting reliable isokinetic measurements at the wrist and elbow (17,18,39,40). However, Peak_Ƭ_ and Score_E_ should not be considered redundant, as Peak_Ƭ_ reflects a single maximal value whereas Score_E_ incorporates torque deviations across the full profile. This is clinically relevant because two patients may present similar peak torque values while differing substantially in the portion of the range of motion over which abnormal resistance is present.

In contrast, MAS agreement was consistently lower, and varied according to the joint and type of comparison. This finding is in line with the known limitations of the MAS, particularly its dependence on examiner perception, manually imposed stretch velocity, and interpretation of ordinal categories (11–13,41). Recent data from Vidmar et al. showed good-to-excellent within-rater reliability for most upper-limb muscle groups but substantially more variable inter-rater agreement, reinforcing the importance of examiner standardization (11). In our study, both inter-rater and test-retest agreement at the elbow were broadly consistent with previous reports (11,41). However, test-retest agreement should be interpreted differently from conventional intra-rater agreement, because V1 value incorporated ratings from both examiners, whereas V2 was performed by one of these examiners. At the wrist, agreement was poorer overall than previously reported in some studies (11,41). This may reflect the greater difficulty of manually grading resistance in a smaller joint with lower torque amplitudes and greater sensitivity to wrist, hand and finger positioning. More broadly, this interpretation is consistent with previous concerns regarding the ambiguity of MAS categories, particularly around grades 1 and 1+, and with evidence that MAS reliability varies according to the tested limb and muscle group (13).

Reliability was generally maintained in both dynamometer testing conditions, although values tended to be slightly higher or more consistent in the ergo chair condition. This may be explained by better proximal stabilization and limb support when patients were seated on the dedicated ergo chair. In the wheelchair condition, the absence of an armrest (Fig 2) may have reduced limb stabilization and relaxation, potentially increasing measurement variability or leading to lower Score_E_ values (Supplementals Fig 4). Nevertheless, reliability remained good in the wheelchair condition, suggesting that this more clinically accessible setup may be acceptable when transfer to the ergo chair is not feasible, provided that positioning is carefully standardized.

### Relationship with MAS and clinical interpretation

Although isokinetic dynamometry is a well-established method for recording joint torque and position during standardized movement, the validity of dynamometer-derived outcomes for assessing post-stroke hyper-resistance depends strongly on the metric used to characterize the torque-angle response. In the present study, the MAS was used as a clinical comparator rather than as a criterion standard, because it remains the most widely used clinical scale while providing only an ordinal and examiner-dependent estimate of resistance to passive movement (11–13).

Moderate positive associations were found between Score_E_ and MAS grades, particularly at the elbow, where significant correlations were observed in both the ergo chair and wheelchair conditions. At the wrist, the association between Score_E_ and MAS was significant in the ergo chair condition but not in the wheelchair condition. This pattern suggests that Score_E_ captures clinically meaningful features of hyper-resistance, while also providing information that is not fully represented by the MAS. Indeed, the dispersion of Score_E_ values within adjacent MAS grades, especially around grades 1 and 1+, suggests that patients assigned to similar MAS grades may nevertheless present substantially different torque-angle profiles. This observation is consistent with previous concerns regarding the subjectivity of the MAS and the ambiguity between grades 1 and 1+ (13). It is also consistent with the broader literature showing that correlations between clinical scales and instrumented biomechanical measures vary substantially depending on the selected parameter, joint, and imposed movement characteristics (4,13,42).

In addition to Score_E_ magnitude, the abnormal angular span tended to be greater in higher MAS categories, as reported in Supplementary Table 1. Although this observation was descriptive and was not formally tested, it supports the clinical relevance of the proposed approach, because MAS grading partly depends on the portion of the range of motion over which resistance is perceived. Thus, the reference-based torque-profile analysis not only quantifies the magnitude of abnormal resistance, but also captures a spatial feature of the response that is embedded in the clinical grading logic of the MAS. The weaker association observed at the wrist, particularly in the wheelchair condition, should be interpreted cautiously. Wrist flexor assessment involves lower torque amplitudes than larger joints and the absence of an armrest and reduced standardization of proximal limb support in the wheelchair condition may have further increased variability in the torque profile. In addition, the narrower distribution of MAS grades at the wrist in this study may have limited the ability to detect associations between Score_E_ and MAS. The lower inter-rater agreement of MAS itself at the wrist may also have attenuated its association with the instrumented measures.

Previous studies have reported associations between Ashworth or MAS scores and instrumented measures such as peak torque, work, stiffness, or threshold angle, particularly in larger joints or lower-limb assessments (14–16,43). However, these associations vary across populations, joints, velocities, and analytical methods. In the present study, Peakτ showed significant associations with MAS only in selected conditions, whereas Score_E_ showed more consistent associations. Taken together, these findings support convergent evidence between Score_E_ and the clinical assessment of passive resistance, while emphasizing that the MAS should not be considered a gold standard for instrumented validation (42).

### ScoreE originality and future perspectives

The main originality of Score_E_ lies in its reference-based analysis of the full torque-angle profile. In contrast to single-point metrics such as Peakτ, Score_E_ quantifies all abnormal samples outside the healthy reference corridor and therefore integrates both the magnitude and the angular distribution of abnormal resistance throughout the movement. Beyond its quantitative value, Score_E_ also provides a visually interpretable representation of the patient’s torque profile relative to healthy reference data, which may facilitate communication between clinicians, engineers, and researchers (4). Unlike existing normative approaches such as NeuroFlexor, the present method compares the patient and reference torque profiles point-by-point across the entire joint excursion, thereby preserving the angular distribution of the deviation (20).

The exploratory PRE–POST vibration analysis did not allow formal assessment of responsiveness, because the intervention did not induce a sufficiently large or consistent modification of hyper-resistance. Although vibration has been reported to reduce post-stroke spasticity in some studies, effects vary across protocols, and in the present study, the 10-minute duration may not have been adequate (44). Responsiveness refers to the ability of an instrument to detect change over time in the construct being measured, therefore, it can only be meaningfully evaluated when the intervention induces a change expected to exceed the measurement variability (45). The present vibration protocol did not provide such a condition. Nevertheless, the continuous distribution of Score_E_ changes illustrates its potential to describe individual variations more finely than an ordinal clinical scale. Future studies should evaluate responsiveness using interventions expected to induce larger, controlled, and clinically meaningful changes. Methodological developments should also include multi-velocity assessments and electromyographic recordings, notably to distinguish neural from non-neural contributors, age-matched normative datasets, anthropometric normalization, and transfer of the torque-profile approach to other joints and more portable or robotic devices (8).

### Study limitations

Several limitations should be acknowledged. First, the reference corridor was built using data from healthy participants who were younger than the post-stroke group. Since passive joint resistance may increase with age, this may have led to an overestimation of Score_E_ in some patients (20,46,47). Reassuringly, illustrative examples (Supplementals Fig 2) showed that torque profiles from limbs rated as MAS = 0, including non-paretic limbs, could remain within the reference corridor and yield Score_E_ = 0. However, these examples do not replace the need for age-matched normative data. Importantly, the use of the same reference corridor across sessions and examiners is less likely to affect reliability estimates, but it may influence the absolute magnitude and clinical interpretation of Score_E_. Future studies should therefore establish larger normative datasets stratified by age.

Secondly, torque signals were analyzed without normalization to limb segment mass and anthropometrics. Although initial offset correction was applied to align each patient profile with the center of the corresponding reference corridor at movement onset, the absence of such normalization may have influenced absolute Score_E_ values and between patients’ comparisons. This limitation should be addressed in future implementations to improve the biomechanical interpretability of Score_E_.

Third, the study population mainly included patients with non-painful, mild-to-moderate hyper-resistance. This was necessary to perform standardized passive mobilization under acceptable testing conditions, but it limits the generalizability of the results to patients with severe hyper-resistance, marked contractures, and painful joints.

Fourth, the identity of the examiner who performed the V2 PRE assessment was either one of the examiners of V1. The mean of the two V1 examiner measurements was therefore used as the V1 reference. Consequently, the test-retest estimates reflect between-visit measurement stability but do not allow temporal variability to be fully separated from potential examiner-related variability. Similarly, test–retest MAS agreement should not be interpreted as strict intra-rater agreement.

Finally, the velocity of passive mobilization during MAS assessment was not controlled and may have varied between examiners. Because resistance to passive stretch and its neural contribution can depend on stretch velocity, this may have introduced additional variability in the correlations between MAS grades and dynamometer-derived metrics.

### Conclusion

This proof-of-principle study supports the feasibility and reliability of a normative torque-profile approach for quantifying elbow and wrist flexor hyper-resistance after stroke. The main novelty of Score_E_ lies in its ability to compare each patient’s torque-angle profile with a healthy reference corridor, thereby quantifying abnormal torque deviation while preserving information about the angular span over which hyper-resistance occurs. This provides a more objective, detailed and visually interpretable assessment than conventional single-point torque metrics or ordinal clinical scales.

Score_E_ showed good to excellent inter-rater and test-retest reliability and moderate convergence with MAS grades, while capturing continuous variations within and across clinical categories. Although responsiveness could not be formally assessed in the present study, this approach appears promising for longitudinal monitoring of targeted joints and for future studies evaluating treatment effects. Further validation using age-matched normative datasets, multi-velocity protocols, electromyographic recordings, and interventions expected to induce clinically meaningful changes will be required before broader clinical implementation.

As rehabilitation moves toward more personalized and data-driven care, instrumented approaches such as Score_E_ may contribute to a *better assessment for better treatment* strategy, ultimately supporting more precise therapeutic decision-making in post-stroke hyper-resistance management.

## Acknowledgments

The authors thank all the participants for their time, commitment, and kindness. We also thank Corentin Delphin for his assistance with participant assessments, and Nicolas Frenot and Lucie Lordon for their contributions to figure design.

## Author contributions

SJ, JG, CP, DL and IF conceived and designed the study. DB and SJ recruited participants and conducted the experimental visits. SJ and DB collected the data. SJ, JG and LR analyzed the data. IF contributed to statistical design and analyses. SJ drafted the manuscript. All authors critically reviewed and approved the final version of the manuscript.

## Data availability

The datasets generated during the current study are available from the corresponding author on reasonable request.

## Ethics approval and consent to participate

The study protocol was approved by the CPP Ile de France IV (number 2021-A03219-32) and registered at ClinicalTrials.gov (NCT05315726). All participants provided informed consent before participation.

## Declaration of generative AI in scientific writing

ChatGPT (OpenAI) was used to assist with language editing and rephrasing during manuscript preparation. All AI-assisted text was critically reviewed by the authors, who take full responsibility for the final content.

