## Supplementals for "A normative torque-profile approach for quantifying elbow and wrist flexor hyper-resistance after stroke: a proof-of-principle study"

### Supplementals material

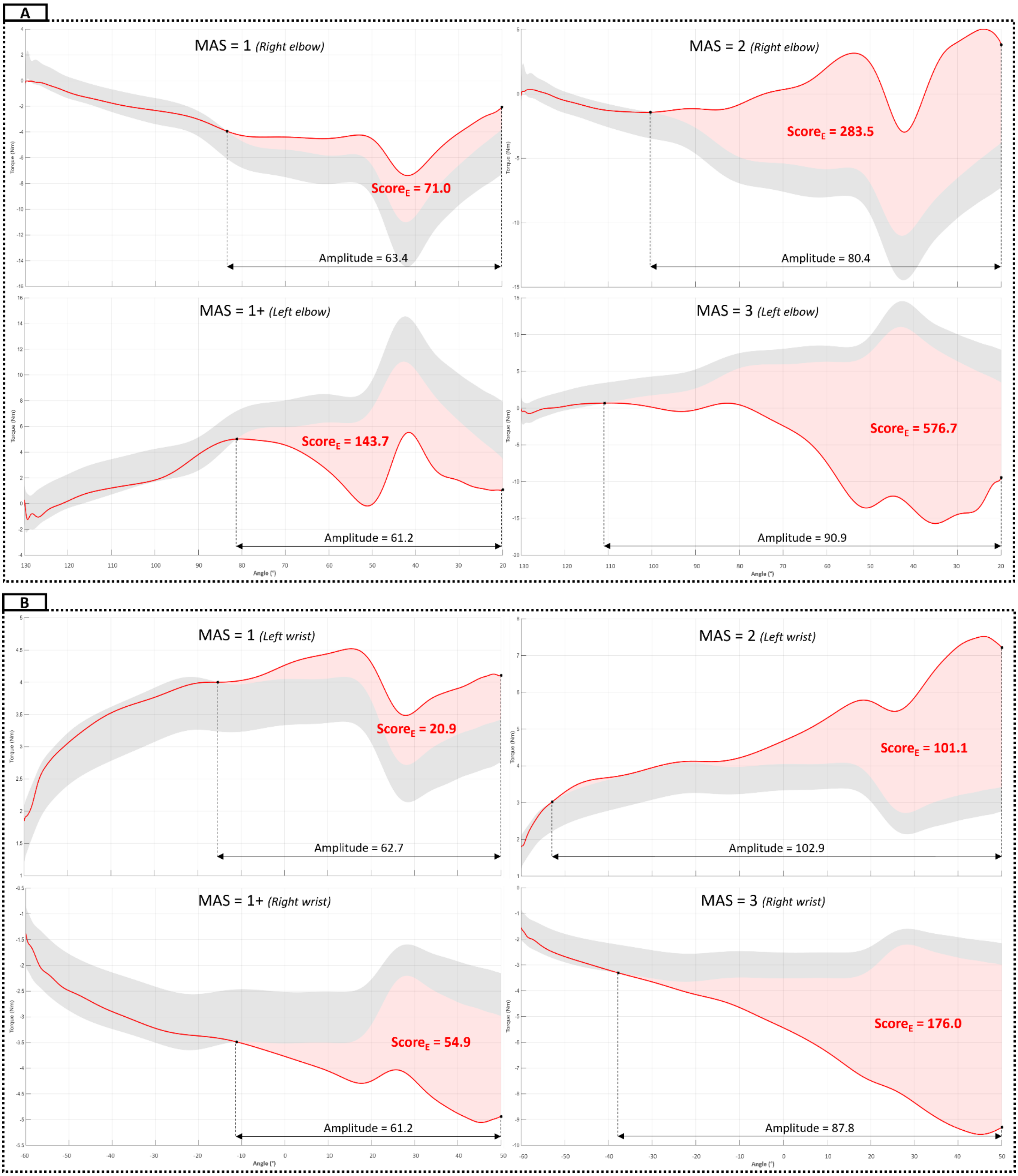


*Suppl Fig 1: Representative $Score_E$ examples across MAS grades. Representative torque-angle profiles obtained in the ergo chair condition are shown for different MAS grades at the elbow (A) and wrist (B). The gray maxarea represents the corresponding healthy reference corridor, the red curve represents the patient's torque profile, and the shaded area indicates samples exceeding the reference corridor and contributing to $Score_E$.*

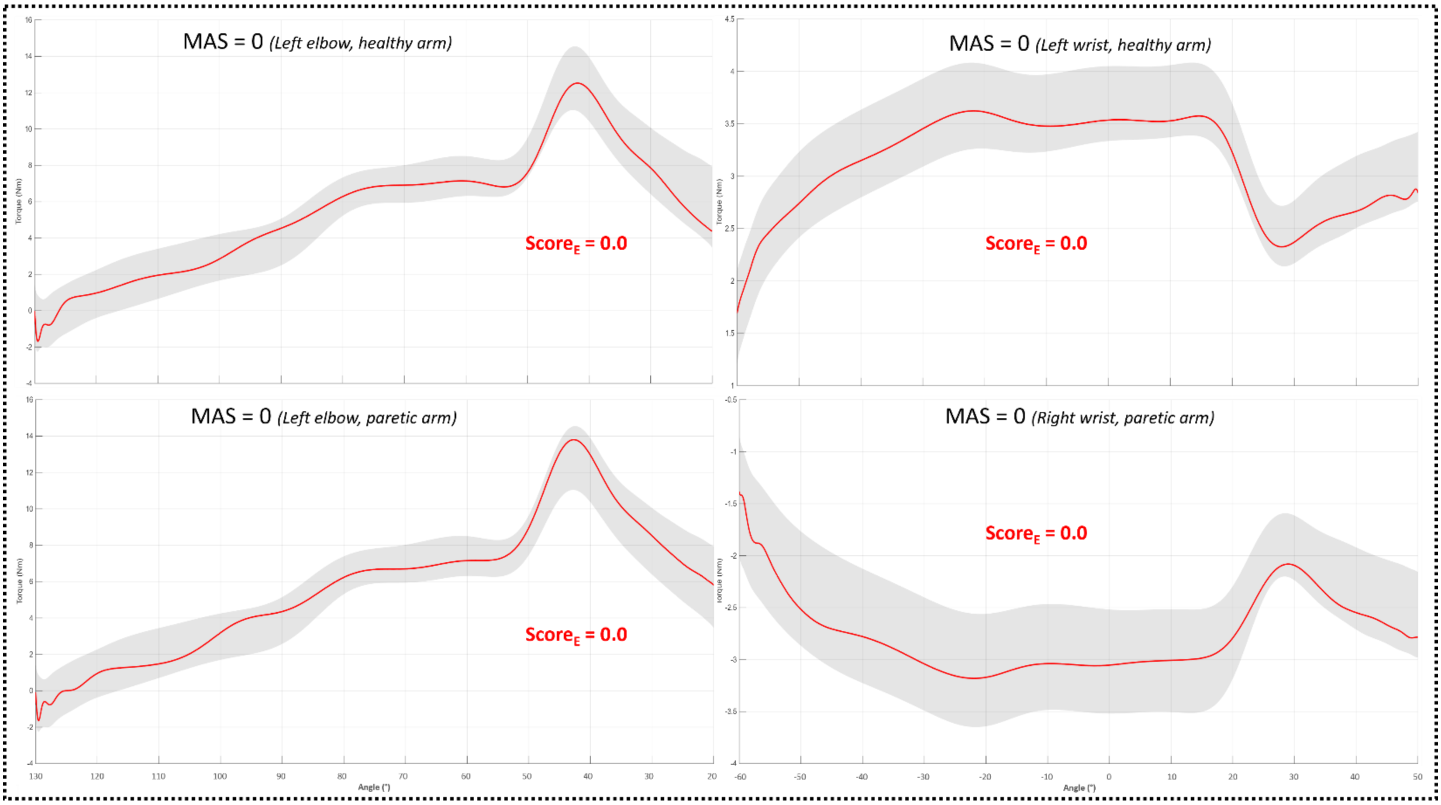


*Suppl Fig 2: Examples of Score$_E$ equal to zero in healthy and paretic limbs. Representative torque-angle profiles are shown for a patient's healthy limb and a paretic limb without spasticity rated as MAS = 0. In each example, the torque profile remained within the corresponding reference corridor, resulting in Score$_E$ = 0.*

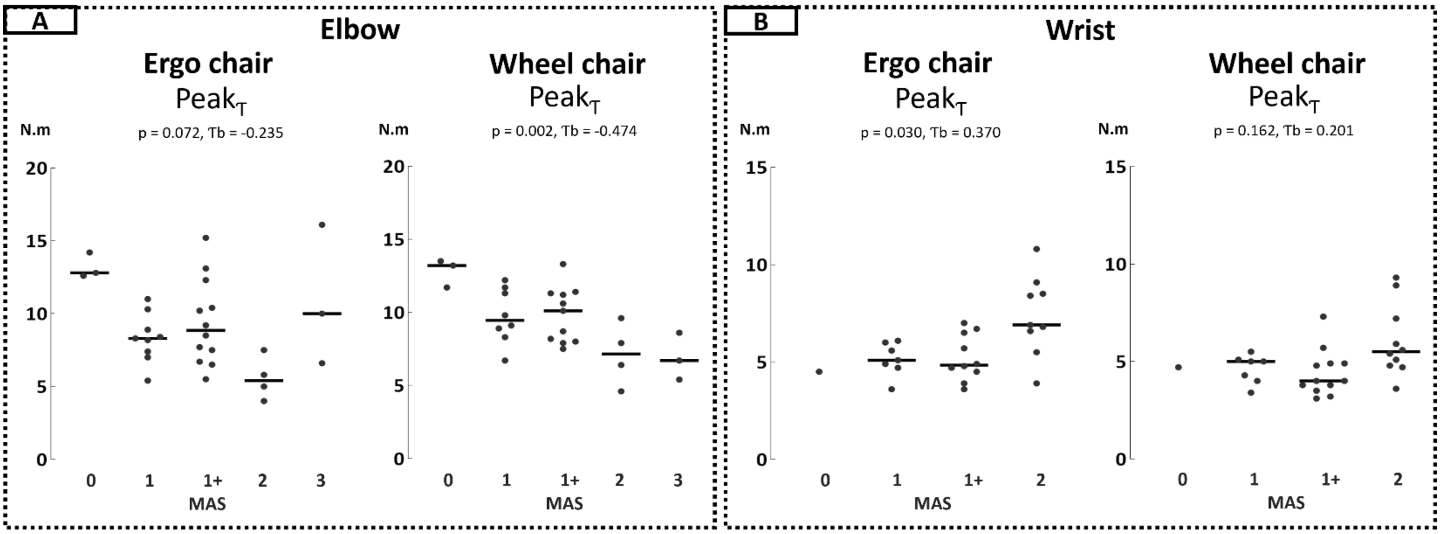


*Suppl Fig 3: Distribution of Peak$_T$ according to MAS grade. Individual Peak$_T$ values are displayed according to MAS grade for elbow (A) and wrist (B) flexors, in the ergo chair and wheelchair conditions. Each dot represents one patient, and horizontal lines indicate median values within each MAS grade. This figure complements Fig. 5 by showing the corresponding distribution for the conventional peak torque metric.*

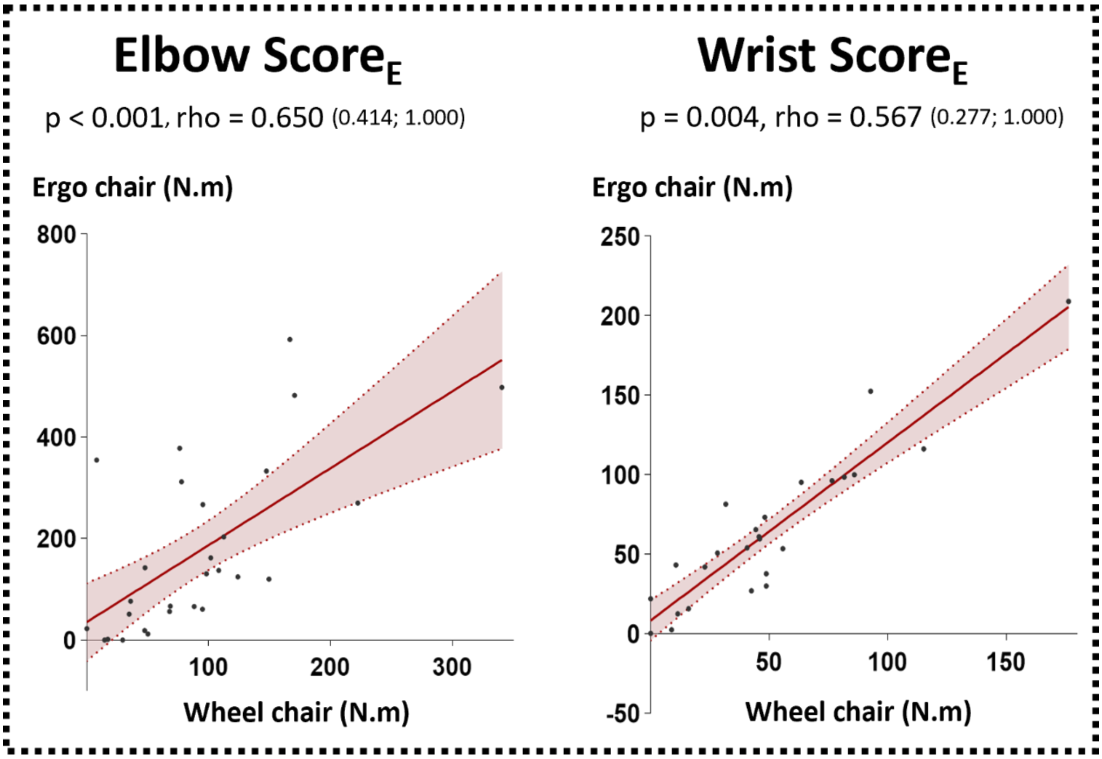


*Suppl Fig 4:* *Association between $Score_E$ values obtained in the ergo chair and the wheelchair conditions. Individual $Score_E$ values obtained in the ergo chair condition are plotted against those obtained in the wheelchair condition for elbow and wrist flexors. Each dot represents one patient. Spearman's rank correlation coefficients (rho), 95% confidence intervals, and corresponding p-values are reported for each joint.*

| Elbow Ergo chair | | | Wrist Ergo chair | | |
|---|---|---|---|---|---|
| MAS | $Score_E$ | Amplitude | MAS | $Score_E$ | Amplitude |
| 0 (n = 3) | 12.3 [10.6] | 42.3 [28.5] | 0 (n = 1) | 27.2 [0.0] | 45.6 [0.0] |
| 1 (n = 9) | 66.4 [74.0] | 64.5 [17.2] | 1 (n = 7) | 42.2 [29.3] | 77.6 [44.6] |
| 1+ (n = 12) | 139.8 [292.0] | 69.7 [30.3] | 1+ (n = 10) | 52.5 [58.4] | 62.7 [25.5] |
| 2 (n = 4) | 236.6 [100.6] | 95.9 [18.3] | 2 (n = 9) | 100.0 [65.4] | 83.1 [30.4] |
| 3 (n = 3) | 482.2 [162.7] | 85.9 [25.8] | 3 (n = 0) | N/A | N/A |

*Suppl Table 1:* *Descriptive $Score_E$ and amplitude values according to MAS grade. Values are presented as median [IQR] for each MAS grade. MAS categories were derived from the mean of the two V1 examiner ratings, with grade 1+ coded as 1.5, using the predefined intervals described in the Methods. $Score_E$ represents the discrete norm of Torque excess and amplitude is expressed in degrees and corresponds to the angular portion of the movement in which the torque profile exceeded the reference corridor.*

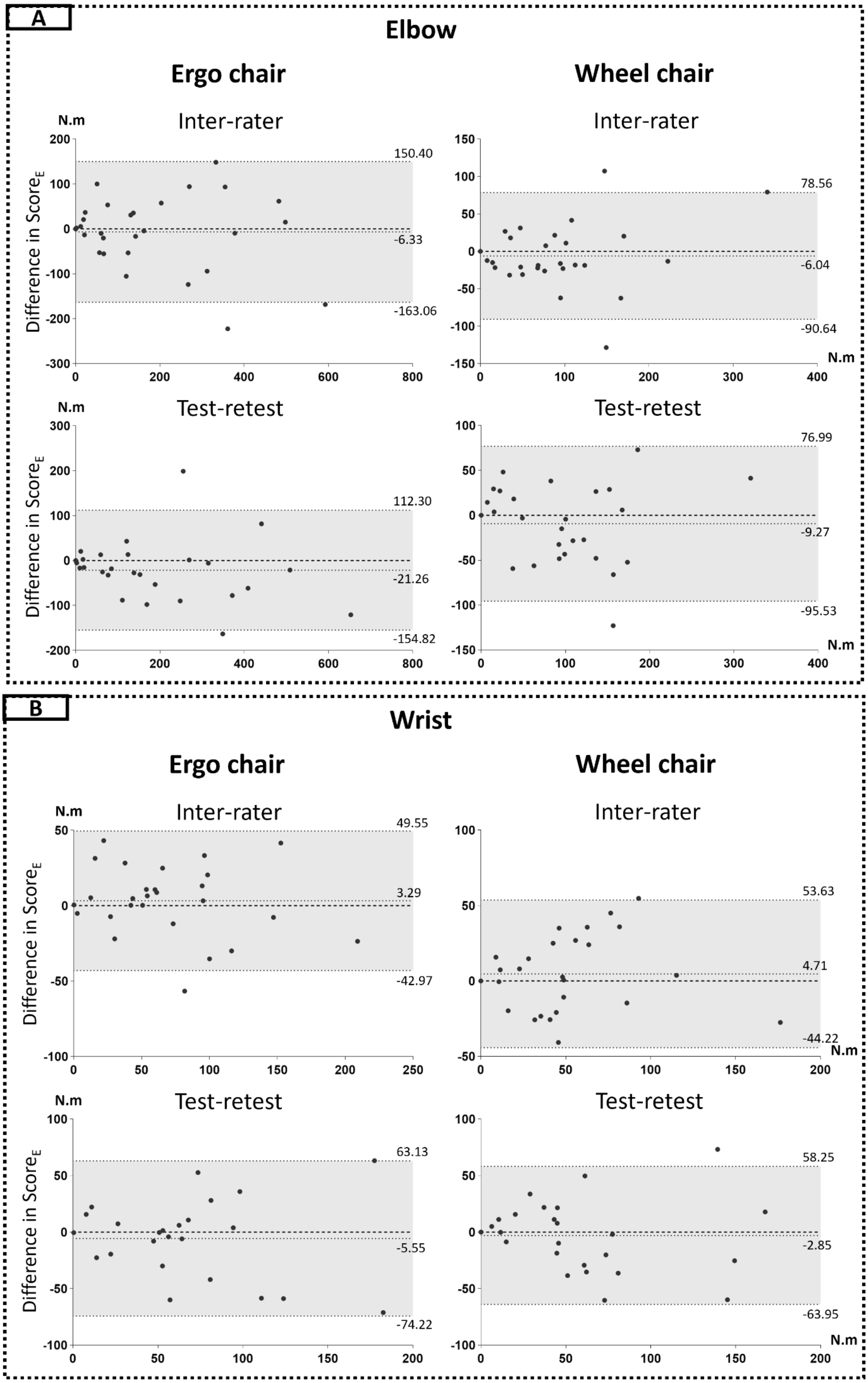


*Suppl Figure 5: Bland-Altman Plots illustrating inter-rater and test-retest agreement for Score$_E$ at the elbow (**Figure A**) and wrist (**Figure B**). Middle lines represent the zero difference and the mean bias (with value) respectively, and dashed lines above and under the grey area indicate the 95% limits of agreement (with values).*
